# High risk of hypoxemic COVID-19 pneumonia in myasthenia gravis patients with type I IFN autoantibodies

**DOI:** 10.64898/2026.03.27.26349525

**Authors:** Adrian Gervais, Astrid Marchal, Alexis Maillard, Tom Le Voyer, Jérémie Rosain, Quentin Philipot, Lucy Bizien, Jessica Peel, Axel Cederholm, Mélanie Migaud, Sylvie Pons, Kahina Saker, Pascal Laforet, Mélodie Aubart, Cyril Gitiaux, Catherine Biggs, Rafael Leon Lopez, Sarah Souvannanorath, Céline Tard, Aleksandra Nadaj Pakleza, Aude-Marie Grapperon, Nicholas Heming, Djillali Annane, Annie Verschueren, Shahram Attarian, Kévin Bigaut, Karolina Hankiewicz, Ludivine Kouton, Rocio-Nur Villar-Quiles, Cécile Cauquil, Marie-Céline Fleury, Emilie Rocher, Guillaume Nicolas, Eduardo de Paula Estephan, Maria da Penha Ananias Morita, Edmar Zanoteli, Zakaria Saied, Amine Rachdi, Amouri Rim, Samir Belal, Samia Ben Sassi, Annemarie Hübers, Emmanuel Faure, Isabelle Desguerre, Clémence Basse, Nicolas Girard, Vivien Béziat, Qiang Pan-Hammarström, Lennart Hammarström, Aaron Bodansky, Audrey V. Parent, Mark S. Anderson, Joseph L. DeRisi, Sophie Demeret, Frédérique Truffault, Romain Fort, Florence Ader, Florent Wallet, Laurent Abel, Thierry Molina, Marie-Alexandra Alyanakian, Rozen Le Panse, Guilhem Solé, Aurélie Cobat, Nils Landegren, Jean-Laurent Casanova, Anne Puel, Paul Bastard, Emmanuelle Jouanguy

## Abstract

Patients with myasthenia gravis (MG) may produce autoantibodies neutralizing type I interferons (AAN-I-IFN), which underlie severe viral diseases, including critical COVID-19 pneumonia, in patients without MG. We studied an international cohort of 85 unvaccinated SARS-CoV-2-infected MG patients not given antiviral treatment. Hypoxemic pneumonia occurred in 48 of these patients, including 22 (45.8%) with AAN-I-IFN, which neutralized both IFN-α2 and IFN-ω in 14 (29.2%) patients. Six (16.2%) of the remaining 37 patients had AAN-I-IFN, neutralizing both IFN-α2 and IFN-ω in three patients. The risk of hypoxemic pneumonia was greater in MG patients with AAN-I-IFN neutralizing 10 ng/mL of both IFN-α2 and IFN-ω (odds ratio and 95% confidence interval (OR [95% CI]): 12.7 [2.1-78.9], *p*=0. 0010) or IFN-α2 at any dose (OR [95% CI]: 4.7 [1.5-15.0], *p*=0.0054) than in those without such autoantibodies. The risk of producing AAN-I-IFN was much higher in MG patients than in the general population (OR [95% CI]: 28.9 [10.8-77.7], *p*=4.9×10^-27^). Thymoma was found in 14 patients and increased the risk of AAN-I-IFN (64% versus 27%, (OR [95% CI]: 5.6 [1.6-19.4], *p*=0.0050) and hypoxemic pneumonia (9.2 [1.9-44.2]; p=0.0019). Thymoma is, thus, associated with a higher risk of producing AAN-I-IFN, which are, in turn, associated with a higher risk of developing life-threatening COVID-19 pneumonia in patients with MG.

**Significance Statement:** Patients with myasthenia gravis (MG) have a higher risk of developing life-threatening COVID-19 pneumonia. In an international cohort of unvaccinated, untreated SARS-CoV-2-infected MG patients, we found that autoantibodies neutralizing type I interferons (AAN-I-IFN) were strongly associated with severe hypoxemic pneumonia. Thymoma further increased the risk of both AAN-I-IFN and severe disease. These findings suggest the existence of a critical immunological mechanism that could guide risk stratification and targeted interventions for MG patients during viral infections, such as COVID-19.

## Introduction

Myasthenia gravis (MG) is an autoimmune disease characterized by muscle weakness, primarily affecting the extrinsic ocular muscles, but also the limb and axial muscles in cases of generalized MG (*1*). The clinical manifestations of MG result from the production of autoantibodies (auto-Abs) targeting acetylcholine receptors (AChR) in about 85%, muscle-specific kinase (MuSK) receptors in about 6%, and/or low-density lipoprotein receptor-related protein 4 (LRP4) in about 2% of MG patients (*2*). MG has a prevalence of 1-2/10,000 individuals (*3*) with a bimodal age distribution: an early-onset peak at about 30 years of age (early-onset MG, EOMG) and a later-onset peak after the age of 50 years (late-onset MG, LOMG) (*3*). Patients with very late-onset MG (after the age of 65 years) appear to be more likely to die from infection than those diagnosed before the age of 65 years (*4*). Treatments include acetylcholinesterase inhibitors, and immunomodulating or immunosuppressive therapies (IST). Thymectomy can also improve the course of MG (*5*). Indeed, thymic abnormalities are common in MG, with about 65% of patients displaying thymic hyperplasia (characterized by an inflamed thymus due to the overproliferation of thymic epithelial cells or hyperplastic lymph follicles (*6, 7*)), and about 15% of MG patients are also diagnosed with thymoma (*8–10*). The pathogenesis of MG remains unclear. MG is thought to have a genetic origin, based on reports of multiplex families (*11*). Genome-wide association studies (GWAS) and HLA association studies have revealed modest effects of common variants (*12, 13*), including signals in the genes encoding Ach-R subunits (*14*).

Auto-Abs neutralizing type I interferons (AAN-I-IFN) have been reported in about 30% of MG patients, up to 70% of patients with MG associated with thymoma, and about 40% of patients with thymoma only (*15, 16*). AAN-I-IFN can be due to monogenic inborn errors of immunity, in patients with inborn errors of central tolerance, for example (*17–25*) (Casanova, JHI 2026, in press). Since 2020, AAN-I-IFN have been shown to be strong determinants of severe COVID-19 detected in about 15% of cases of life-threatening COVID-19 pneumonia (*26*), including cases in children (*27*), 20% of all fatal COVID-19 cases tested (*28, 29*), and about 20% of individuals suffering from ‘breakthrough’ hypoxemic COVID-19 pneumonia despite a normal Ab response to mRNA vaccination (*30, 31*). These auto-Abs are thought to persist throughout the individual’s lifetime (*32*) and have also been detected in 5% of cases of critical seasonal influenza pneumonia (*33*), a fatal case of H5N1 avian influenza (*34*), 38% of cases of herpetic fulminant hepatitis (*35*), 25% of cases of critical Middle East respiratory syndrome (MERS) pneumonia (*36*), 40% of cases of West Nile virus encephalitis (*37, 38*), 10% of severe tick-borne encephalitis cases (*39*), in cases of the rarer, severe Powassan virus encephalitis, Usutu virus encephalitis, and severe Ross River virus disease (*40*), and one third and 60%, respectively, of severe adverse reactions to yellow fever YFV-17D and chikungunya VLA1553 live-attenuated viral vaccines (*41, 42*). Importantly, it has been established that germinal center-derived memory B-cell responses directed against type I IFNs are present before severe viral infection (*43*). In a French cohort of 3,558 MG patients, 0.96% suffered SARS-CoV-2 infection before vaccination, with about 30% of these patients developing life-threatening COVID-19 pneumonia (*44*). In a Canadian cohort of 4,411 MG patients, 3.7% contracted COVID-19, including 30.5% who were hospitalized (*45*). We, thus, hypothesized that AAN-I-IFN may underlie life-threatening COVID-19 in patients with MG.

## Results

### AAN-I-IFN in an independent pre-COVID-19 cohort of 86 MG patients

We analyzed a cohort of MG patients from whom blood samples had been collected before the emergence of COVID-19. The median age of these patients was 63 years (range: 14-88 years) and 41 (48%) were men. No information about comorbid conditions, thymoma, thymectomy or treatments was available for this cohort (Table 1). We used an automated, sensitive, ELISA-like technique (Gyros) to measure the levels of circulating auto-Abs against IFN-α2 or IFN-ω in the plasma or serum of all MG patients, as previously described (*28*). In total, 29 (34%) of the 86 MG patients had intermediate (between 30 and 100 arbitrary units (AU)) or high (>100 AU) levels of auto-Abs against IFN-α2 and/or IFN-ω, consistent with previous reports (*16*) (Figure 1A). The auto-Abs recognized IFN-α2 only (76%), IFN-ω only (7%), or both (17%). Three patients (3%) also had auto-Abs against IFN-β, as detected by ELISA (Figure 1B). We then investigated the ability of these antibodies to neutralize type I IFNs in a luciferase reporter assay, as previously described (*28*). Remarkably, 37 (43%) patients had AAN-I-IFN at a concentration of 100 pg/mL: neutralizing IFN-α2 only in 7 of 86 (8%), IFN-ω only in 4 (5%), and both IFN-α2 and IFN-ω in 26 (30%) (Figure 1C-D). AAN-I-IFN neutralizing type I IFNs at a concentration of 10 ng/mL were found in 24 (28%) patients: in 7 (8%) patients, they neutralized IFN-α2 only, in 3 (3%), IFN-ω only, and in 14 (16%), both IFN-α2 and IFN-ω, this last group including one patient whose auto-Abs also neutralized IFN-β (Figure S1A). Neutralization was mediated by immunoglobulins (Ig)-G (Figure 1E). Age, sex, and MG serotype were not correlated with AAN-I-IFN production (Figure S1B-F). AAN-I-IFN were strongly associated with MG, even after Bonferroni correction for eight combinations tested (*p*-value from 4.5×10^-35^ for auto-Abs neutralizing 100 pg/mL of IFN-α2 only, to 3.6×10^-21^ for auto-Abs neutralizing both IFN-α2 and IFN-ω at 10 ng/mL). The OR [95% CI] for the presence of AAN-I-IFN in MG patients relative to the general population ranged from 32.1 [12.3-84.1] for auto-Abs neutralizing 100 pg/mL IFN-α2 or IFN-ω, to 89.1 [28.0-282.8] for auto-Abs neutralizing both IFN-α2 and IFN-ω at 10 ng/mL (Figure 1F). Neutralizing IgG AAN-I-IFN were found in pre-COVID-19 blood samples from 43% of the MG patients in this cohort, consistent with previous reports based on other detection methods. We tested the hypothesis that AAN-I-IFN affected the severity of SARS-CoV-2 infection in MG patients.

**Figure 1:**
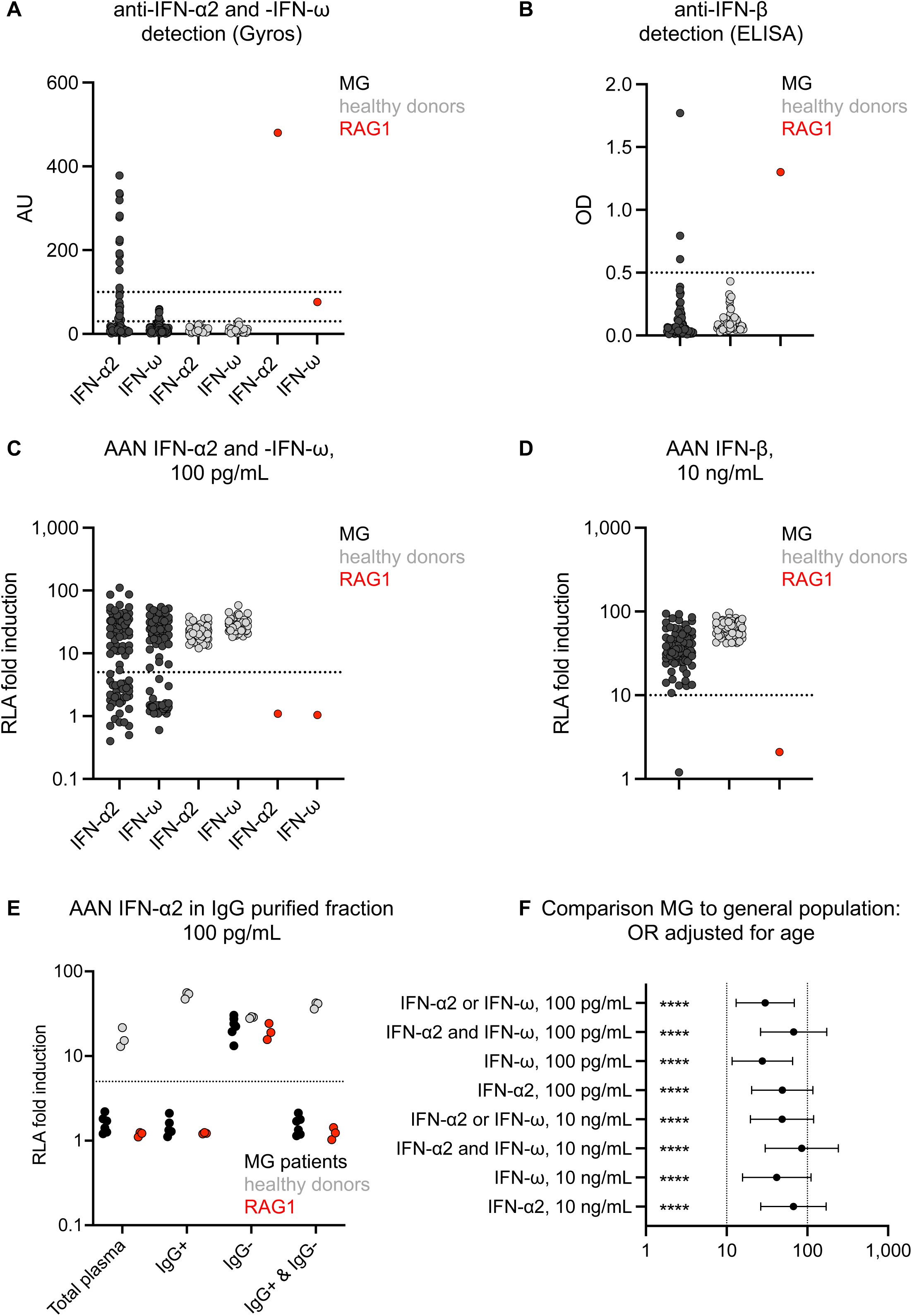
Auto-Abs against type I IFN before COVID-19 in the 86 MG patients. **(A)** Detection of auto-Abs against IFN-α2 and IFN-ω by Gyros. Signals between 30 and 100 AU were considered intermediate, and signals >100 were considered high. RAG1: plasma from a patient with an autosomal recessive (AR) partial recombination activating gene 1 (RAG1) deficiency. **(B)** Detection of auto-Abs against IFN-β by ELISA. Signals >0.5 are considered positive. **(C)** Neutralization of IFN-α2 and IFN-ω at a concentration of 100 pg/mL by auto-Abs in the plasma of MG patients. **(D)** Neutralization of IFN-β at a concentration of 10 ng/mL by auto-Abs in the plasma of MG patients. RAG1: plasma from a patient with AR complete RAG1 deficiency. In panels C and D, RLA fold induction is the stimulated firefly/*Renilla* luminescence ratio normalized against the non-stimulated firefly/*Renilla* luminescence ratio for each individual. **(E)** AAN-IFN-I neutralizing 100 pg/mL IFN-α2 in IgG-enriched and IgG-depleted fractions of plasma. **(F)** Risk of carrying AAN-IFN-I in MG patients relative to that in the general population, with adjustment for age.

### A cohort of 85 patients with myasthenia gravis and COVID-19

Between 2020 and 2022, we recruited an international cohort of 85 unvaccinated MG patients who suffered from SARS-CoV-2 infection and who received no antiviral treatment for COVID-19 (Table 2). Median age was 54 years (23-90 years) and 48% of the patients were men (Table 2). The patients originated from France (*n*=56), Spain (*n*=2), Switzerland (*n*=2), Canada (*n*=1), Brazil (*n*=17), and Tunisia (*n*=7). MG was generalized in 96.4% of patients: 70.6% with AChR auto-Abs, 3.5% with MuSK auto-Abs, 21.2% negative for both these auto-Abs, and 4.7% of unknown status (Table 2). No testing was performed for LPR4 auto-Abs. The most frequent comorbid conditions observed were arterial hypertension (21%) and obesity (13%), often in association. Thymectomy had been performed on 34 (40%) individuals, 14 (16%) of whom had thymoma-associated MG and 20 (24%) of whom had no detectable thymoma. Thymoma was diagnosed by imaging, with post-thymectomy histopathological confirmation, but no further details were available. At the time of SARS-CoV-2 infection, the patients were on the following treatments for MG: acetylcholine esterase inhibitors alone (10%), corticosteroids alone (23.5%), another immunosuppressive therapy alone (10%), or a combination of treatments (56.5%) (Table 2). Overall, 75% of the patients were on at least one immunomodulatory treatment (corticosteroids and/or immunosuppressant) at the time of SARS-CoV-2 infection, and 56% were on at least one B cell-immunosuppressive treatment (azathioprine and/or mycophenolate mofetil and/or rituximab).

### Higher risk of hypoxemic COVID-19 in MG patients with AAN-I-IFN

Patients were classified into two groups — with (*n*=48) and without (*n*=37) hypoxemic COVID-19 pneumonia (Table 3) — and tested for the presence of AAN-I-IFN. We found that 18 of 48 (38%) patients in the hypoxemic group had auto-Ab levels >30 arbitrary units (AU), whereas this was the case for 9 (24%) of the patients in the non-hypoxemic group (*p*=0.21) (Figure 2A). High titers of auto-Abs against IFN-α2 were almost always neutralizing, whereas this was not the case for auto-Abs against IFN-ω (Figure S2A-B). AAN-I-IFN were detected in 22 (46%) of the hypoxemic cases, including one patient (2.1%) with antibodies neutralizing all circulating type I IFNs (i.e. IFN-β, IFN-α2 and IFN-ω), whereas only 6 (16%) of the non-hypoxemic cases had AAN-I-IFN (*p*=0.005, Fisher’s exact test) (Figure 2B-C, Table 4). No major differences were observed between men and women or between age groups (Table 5). AAN-I-IFN neutralized high concentrations (10 ng/mL) of type I IFNs in about two thirds of the patients, and lower concentrations (100 pg/mL) in the remaining third (Figure S2C-D). The risk of hypoxemic pneumonia in MG patients was significantly higher in patients with auto-Abs neutralizing IFN-α2 and/or IFN-ω at a concentration of 10 ng/mL (OR [95% CI]: 12.7 [2.1-78.9], *p*=0.0010), or 100 pg/mL (OR [95% CI]: 3.5 [1.2-10.7], *p*=0.0206), and in those with auto-Abs neutralizing IFN-α2 at any dose (OR [95% CI]: 4.7 [1.5-15.0], *p*=0.0054). Strikingly, 14 of 17 (82%) patients with auto-Abs neutralizing at least 100 pg/mL of both IFN-α2 and IFN-ω had hypoxemic COVID-19 pneumonia (Figure 2D). Overall, AAN-I-IFN neutralizing any concentration of both IFN-α2 and IFN-ω, or IFN-α2 only, were significantly associated with a higher risk of hypoxemic COVID-19 pneumonia in MG patients, even after correction for multiple testing. Adjustment for sex, obesity, and hypertension did not modify this finding.

**Figure 2:**
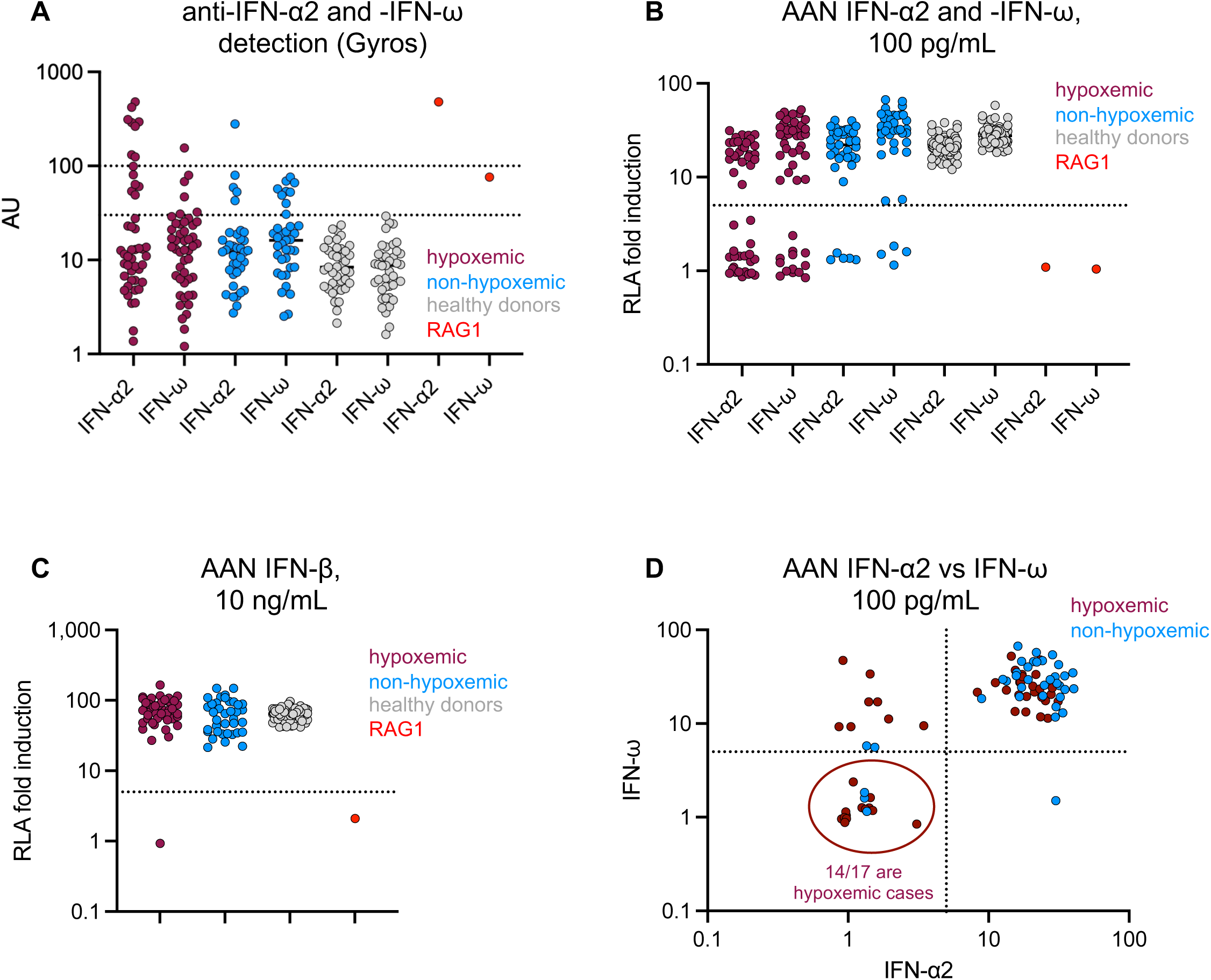
Auto-Abs against type I IFNs in the 85 MG patients infected with SARS-CoV-2. **(A)** Detection of auto-Abs against IFN-α2 and IFN-ω by Gyros, by COVID-19 severity class. **(B)** Neutralization of IFN-α2 and IFN-ω at a concentration of 100 pg/mL by auto-Abs in the plasma of MG patients, by COVID-19 severity class. **(C)** Neutralization of IFN-β at a concentration of 10 ng/mL by auto-Abs in the plasma of MG patients, by COVID-19 severity class. **(D)** Correlation between the neutralization of non-glycosylated IFN-α2 and IFN-ω at a concentration of 100 pg/mL in the hypoxemic and non-hypoxemic groups.

### Higher risk of hypoxemic COVID pneumonia and AAN-I-IFN in MG patients with thymoma

The OR for harboring auto-Abs capable of neutralizing type I IFNs at a concentration of 100 pg/mL in MG patients was 28.9 (95% CI: [10.8-77.7], *p*=4.9×10^-27^) relative to the age-adjusted general population. No significant differences in the prevalence of AAN-I-IFN (*p*=0.6) or hypoxemic COVID-19 (*p*=0.054) were observed between EOMG and LOMG. By contrast, thymoma was strongly associated with the presence of AAN-I-IFN (Figure 3A), consistent with previous reports (*16*). Indeed, AAN-I-IFN were detected in 9 of the 14 MG patients (64%) with thymoma but in only 19 of the 71 (27%) without thymoma (*p*=0.011, Fisher’s exact test). The OR for harboring auto-Abs neutralizing IFN-α2 and/or IFN-ω at a concentration of 100 pg/mL was 5.6 (OR [95% CI]: 1.6-19.4, *p*=0.0050, Firth regression) in MG patients with thymoma relative to those without. Strikingly, 12/14 (86%) patients with thymoma had hypoxemic pneumonia, including seven with auto-Abs neutralizing the higher concentration (10 ng/mL) and one neutralizing the lower concentration (100 pg/mL) of type I IFNs. Seven of these 12 patients (58%) had auto-Abs neutralizing both IFN-α2 and IFN-ω. The OR of hypoxemic pneumonia in MG patients with thymoma relative to those without thymoma, adjusted for age and regardless of the presence or absence of AAN-I-IFN, was 9.2 (OR [95% CI]: 1.9-44.2, *p*=0.0019, Firth regression) (Figure 3B). The observed trend towards a protective effect of thymectomy against hypoxemic COVID-19 in patients without thymoma was not significant (OR [95% CI]: 0.37 [0.1-1.4], *p*=0.13), perhaps due to the small number of patients (Table 6). In patients without detectable AAN-I-IFN, thymoma was associated with hypoxemic COVID-19, suggesting that other forms of autoimmunity, or different pathogenic mechanisms, might contribute to the pathogenesis of severe COVID-19 in these patients. Despite the small number of cases included in this cohort, thymoma was found to be significantly associated with a higher prevalence of AAN-I-IFN and a higher risk of severe COVID-19 in MG patients.

**Figure 3:**
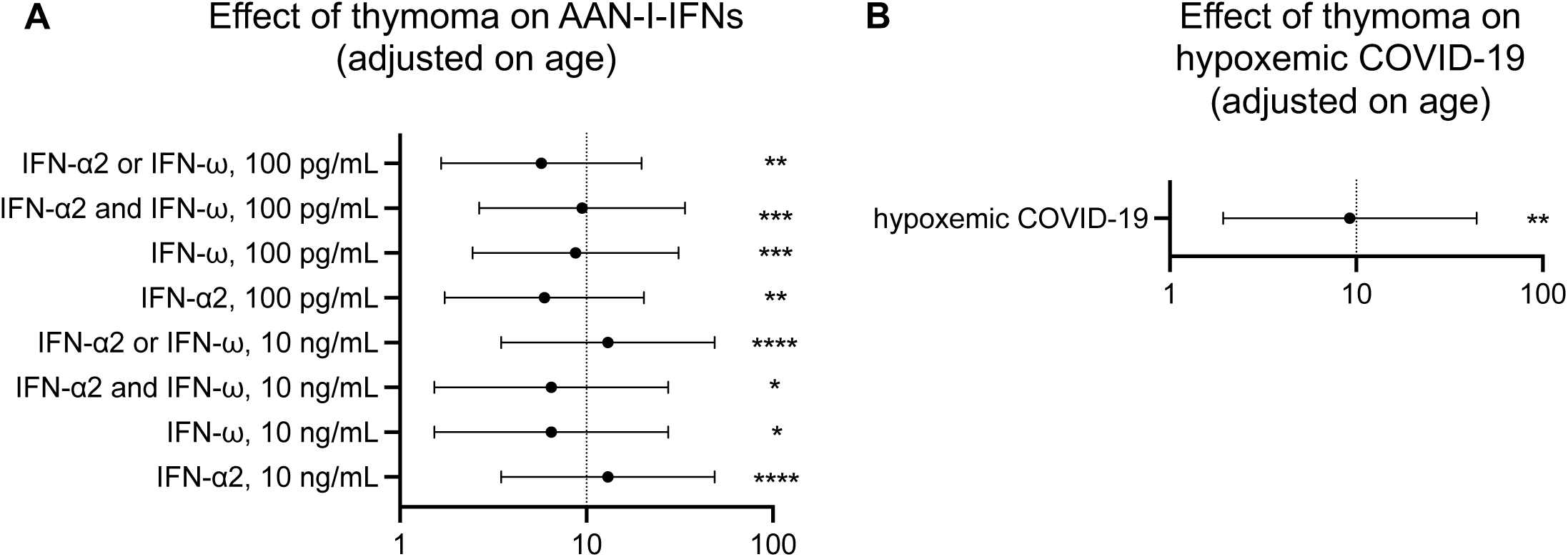
Effects of the presence of thymoma on AAN-I-IFN and hypoxemic COVID-19. **(A)** OR of thymoma for the presence of different combinations of AAN-I-IFN relative to MG patients without thymoma. **(B)** Effect of thymoma on the severity of COVID-19 (hypoxemic disease) relative to that in MG patients without thymoma.

### Neutralization of different type I IFN subtypes and of glycosylated type I IFNs

The 12 IFN-α subtypes display a high degree of sequence identity (70-80%) (*46, 47*). We nevertheless assessed the neutralization of each IFN-α subtype at a concentration of 1 ng/mL^58^. All plasma samples neutralizing IFN-α2 at 10 ng/mL neutralized all IFN-α subtypes (Figure 4A). Conversely, none of the nine plasma samples from MG patients without detectable auto-Abs neutralizing IFN-α2 or IFN-ω tested were found to neutralize any of the other IFN-αs. The single plasma sample neutralizing IFN-α2 only at 100 pg/mL tested was able to neutralize only about half the other IFN-α subtypes at a concentration of 1 ng/mL. The single patient tested with auto-Abs neutralizing both IFN-α2 and IFN-ω at 100 pg/mL had a very similar neutralization pattern, with the neutralization of only certain subtypes of IFN-α, at a concentration 1 ng/mL. These observations may reflect slight differences in concentrations or activity between the IFN-α subtypes tested rather than specificity. Four human type I IFNs (IFN-α2a/b, IFN-α14, IFN-ω, and IFN-β) are normally produced and secreted as glycosylated proteins. We therefore tested the ability of serum samples from all patients to neutralize glycosylated IFN-α2 and IFN-ω at a concentration of 100 pg/mL (we had already tested glycosylated IFN-β in the initial experiment). In a previous study, we found no difference in neutralization capacity between glycosylated and unglycosylated forms (*27*). All patient samples neutralizing non-glycosylated IFN-α or IFN-ω also neutralized their glycosylated forms. Interestingly, the samples of one patient neutralized glycosylated but not non-glycosylated IFN-α2, whereas those of four others neutralized glycosylated but not non-glycosylated IFN-ω (Figure 4B-C), including those of one patient in the hypoxemic group that did not neutralize any other type I IFNs. This observation may reflect a biochemical reality or result from an artifact caused by slight differences in concentration or activity between glycosylated and non-glycosylated IFN-I. Auto-Abs against IFN-α2 generally neutralize all IFN-α subtypes, and they may have a higher affinity or avidity for the glycosylated, physiological form of type I IFNs.

**Figure 4:**
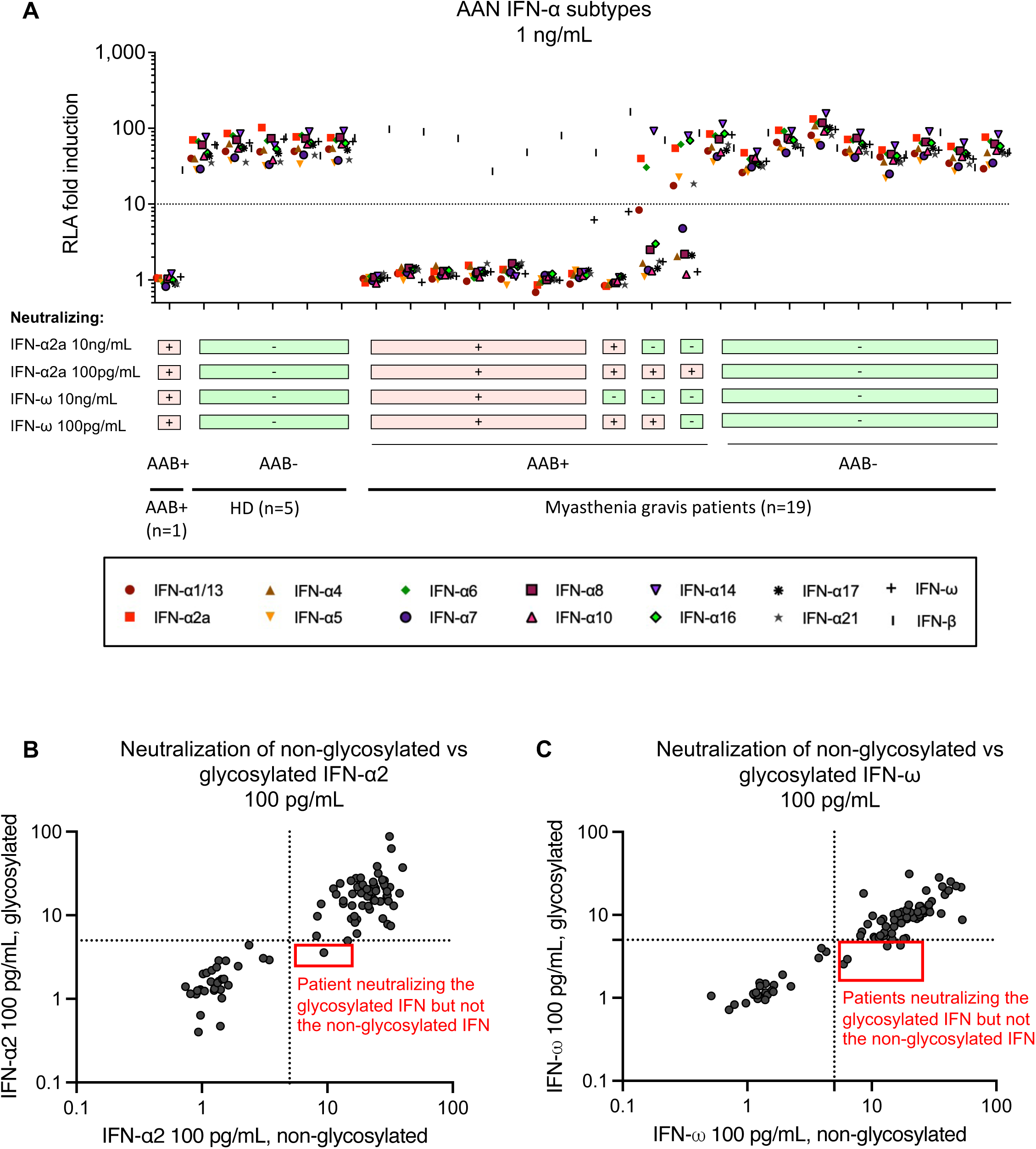
Characterization of auto-Ab-mediated type I IFN neutralization. **(A)** Neutralization of all 12 IFN-α subtypes at a concentration of 1 ng/mL (luciferase) by the auto-Abs of a subset of MG patients. **(B)** Correlation between the neutralization of 100 pg/mL IFN-α2 and that of the same concentration of glycosylated IFN-α2 by the auto-Abs of MG-COVID-19 patients. **(C)** Correlation between the neutralization of 100 pg/mL IFN-ω and that of the same concentration of glycosylated IFN-ω by the auto-Abs of MG-COVID-19 patients.

### Longitudinal samples and impact of immunosuppression, plasmapheresis, or early COVID-19 treatment

For six individuals, we obtained samples collected before and after SARS-CoV-2 infection (Figure 5A). Three of these patients, all with mild disease, had no detectable AAN-I-IFN before or after COVID-19. Another two patients, also with mild disease, had auto-Abs neutralizing IFN-ω at 100 pg/mL before the pandemic, which were no longer detectable after COVID-19 in one of these patients, perhaps secondary to immune suppression. Finally, another patient with hypoxemic disease had auto-Abs neutralizing both IFN-α2 and IFN-ω at 100 pg/mL before the pandemic, with detectable neutralization of IFN-α2 after COVID-19. These findings suggest that AAN-I-IFN may also have been present before infection in the other hypoxemic cases. However, prospective studies based on longitudinal analyses of MG patients are required before any firm conclusions can be drawn. The prevalence of hypoxemic COVID-19 did not differ significantly between MG patients with (*N*=47, 62%) and without (*N*=38, 50%) immunosuppressive therapy (azathioprine + rituximab + MFM) (*p*=0.38). Interestingly, the OR for hypoxemic COVID-19, after adjustment for age, was 6.5 (OR [95% CI]: 0.98-42.6, *p*=0.032, Firth regression) in patients with MG treated with rituximab. This trend, although not significant after correction for multiple testing, is consistent with the results of other studies (*48*). Nevertheless, these findings will require confirmation in larger cohorts. Corticosteroid treatment was not associated with a significantly higher prevalence of auto-Abs or with a poorer COVID-19 outcome (OR 1.6 [0.6-4.3]; *p*=0.33). One patient with hypoxemic pneumonia was treated by plasmapheresis. Despite a relative decrease in IgG anti IFN-α2 levels during successive plasmapheresis sessions, anti-IFN-α2 IgG concentrations remained >913 ng/mL, consistent with sustained serum neutralizing activity against IFN-α2 at 10 ng/mL across all time points (Fig 5B). Finally, another two individuals with AAN-I-IFN (both with auto-Abs neutralizing IFN-α2 at 10 ng/mL, and one with auto-Abs also neutralizing IFN-ω at 100 pg/mL) were treated for COVID-19 early in the course of the disease and were, therefore, analyzed separately from the 85 untreated patients. One received monoclonal Abs (mAbs) against SARS-CoV-2 (bamlanivimab/etesivimab), whereas the other received intravenous dexamethasone. Both had a favorable outcome (non-hypoxemic disease), potentially suggesting that, in patients with MG and AAN-I-IFN, early antiviral and/or anti-inflammatory treatment may mitigate the risk of severe disease, whereas plasmapheresis may not be sufficient to eliminate all the AAN-I-IFN and protect against a poor outcome.

**Figure 5:**
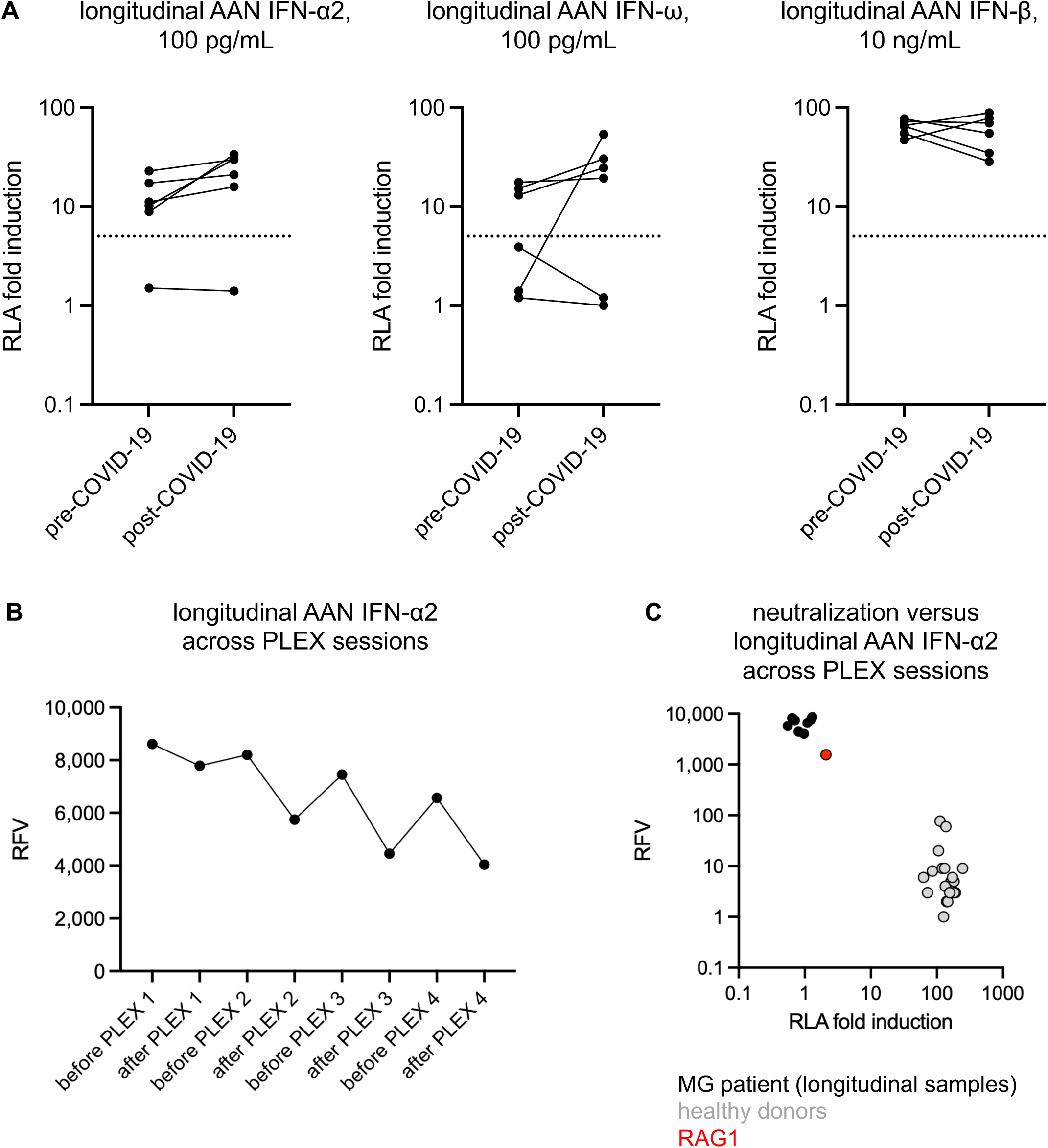
Longitudinal analysis of AAN-I-IFN in six patients sampled before and after the COVID-19 episode, and in a patient treated by plasmapheresis. **(A)** Neutralization of IFN-α2 (left panel), IFN-ω (middle panel) and IFN-β (right panel) in longitudinal samples from 6 MG patients for whom plasma samples obtained before and after COVID-19 episode were available. **(B)** Anti-IFN-α2 IgG titers in eight plasma samples from a patient with hypoxemic COVID-19 pneumonia, before and after four plasmapheresis (PLEX) sessions. Anti-IFN-α2 IgG titers were measured in the VIDAS anti-IFN-α2 IgG prototype assay (bioMérieux, Marcy L’Etoile, France). The titers of all eight samples were above the upper limit of quantification, so relative fluorescence values (RFV) are provided instead of concentrations. **(C)** Anti-IFN-α2 IgG titers (*y*-axis) and neutralization (*x*-axis) in eight plasma samples from a patient with hypoxemic COVID-19 pneumonia, before and after four PLEX sessions. Anti-IFN-α2 IgG titers were measured in the VIDAS anti-IFN-α2 IgG prototype assay (bioMérieux, Marcy L’Etoile, France). Neutralization was assessed with the luciferase assay described above.

### No detectable impact of other auto-Abs on COVID-19 severity

Given the autoimmune nature of MG, we investigated the possibility of broader autoimmunity in these patients. We first performed Luminex® screening for auto-Abs against other cytokines. Some patients had detectable auto-Abs against IFN-γ, IL-27, or TNF-α, but the prevalence of these auto-Abs did not differ significantly between the hypoxemic and non-hypoxemic groups, suggesting that these auto-Abs had no significant impact on COVID-19 severity (Figure S3). The detection of auto-Abs against IFN-α, IFN-ω, and IFN-β in Luminex® assays was well correlated with their detection by Gyros and with the neutralization data (Figure S4). We also used a broader, complementary, unbiased approach to detect additional auto-Abs: proteome-wide auto-antigen microarray (HuProt™) (*49*) for extensive auto-Ab screening. We tested a subset of MG patients with (*n=*11) and without (*n=*2) AAN-I-IFN, including seven patients with thymoma, and 22 healthy controls. In the HuProt™ microarray, a strong signal for auto-Abs against IFN-α subtypes and IFN-ω was observed in the patients testing positive by other means (Figure 6A), whereas the signal was very weak for AAN-I-IFN-negative patients (Figure 6B). Surprisingly, no auto-Abs against AChR or MuSK were detected, even though most of the patients tested are known to carry them, probably because these multimeric or transmembrane proteins that are likely to be incorrectly folded in this screening assay. The lung-specific autoantigens (BPIFB1 and KCNRG) reported in APS-1 and thymoma patients (*50*) were not detected in our cohort. However, strong signals were obtained for several other auto-Abs in several patients (Figure 6C). Five patients (including 4 with thymoma) had very consistent screening results highly suggestive of auto-Abs against KLHL (Kelch-like family) proteins associated with various forms of congenital myopathy (*51, 52*). Overall, the presence of AAN-I-IFN increases the risk of hypoxemic COVID-19 pneumonia, which was not the case for the other auto-Abs detected here.

**Figure 6:**
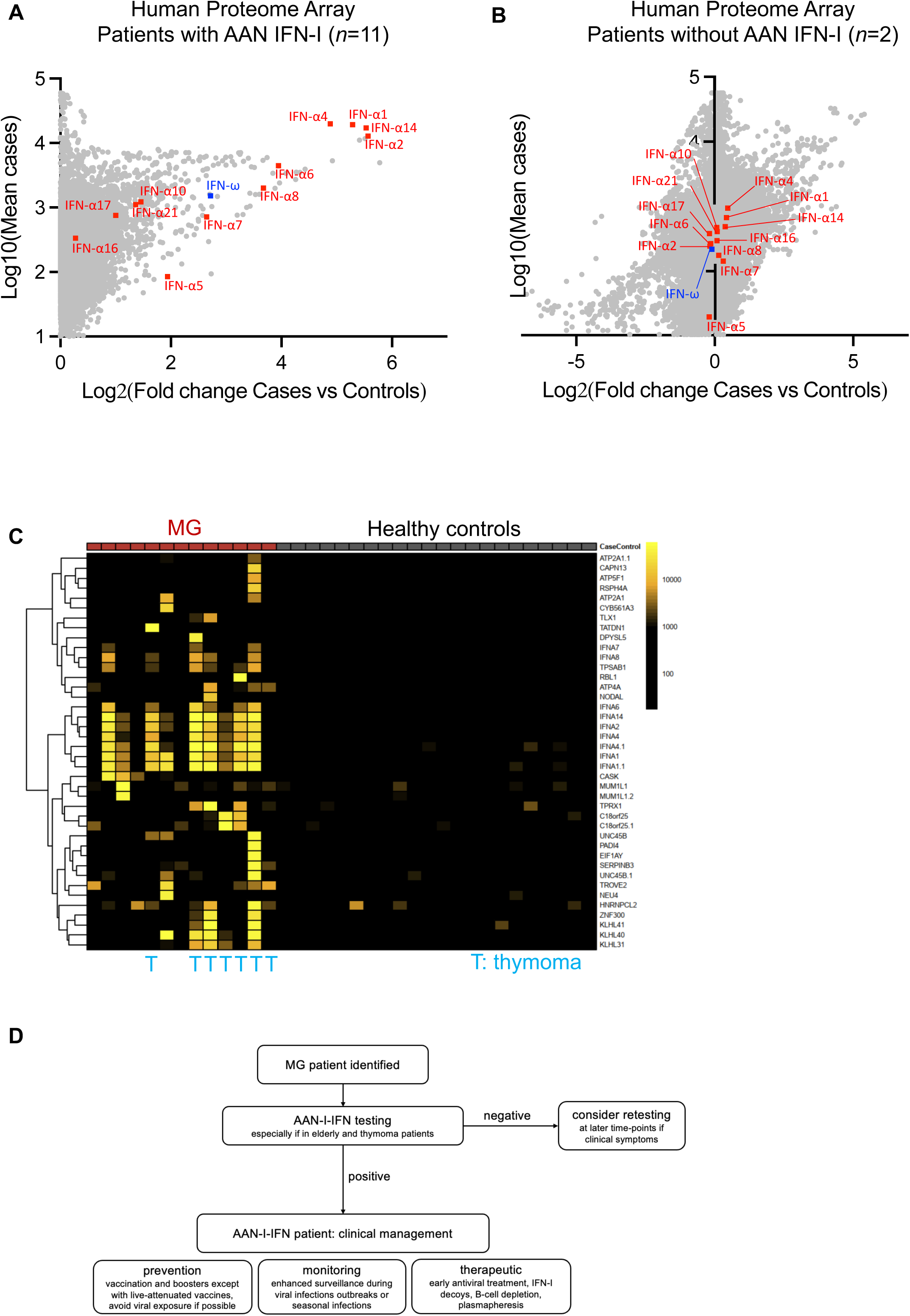
Broad auto-Ab spectrum analysis in a subset of MG patients. **(A)** HuProt® auto-Ab profiling of 11 MG patients with AAN-I-IFN identified in a luciferase assay. Results are displayed as the log_2_ fold-change difference between cases and controls plotted against the log_10_ mean signal for cases. Results for type I IFNs are indicated in red (IFN-αs) or blue (IFN-ω). **(B)** HuProt® auto-Ab profiling of two MG patients without AAN-I-IFN identified in a luciferase assay. **(C)** Heatmap showing the top targets of HuProt® auto-Ab profiling for the 13 MG patients tested. **(D)** Proposed clinical framework for the screening and management of AAN-I-IFN in patients with MG.

## Discussion

We describe a cohort of 85 MG patients infected with SARS-CoV-2 before vaccination. Consistent with prior findings, many of these patients (33%) were found to harbor AAN-I-IFN (*15, 16*). We found that these AAN-I-IFN increased the risk of hypoxemic COVID-19 pneumonia. These auto-Abs were detected before SARS-CoV-2 infection in the cases tested, demonstrating their presence before infection in all MG patients with AAN-I-IFN, as reported in other cohorts (*15, 16*). In some cases, such auto-Abs may be transient and appear during acute viral infection or in other contexts (*53*), but several recent studies suggest that these auto-Abs typically persist, diversify, and increase in potency/concentration (*54, 55*), (Fournier et al., in press). Studies of larger numbers of pre- and post-infection samples would improve characterization of the temporal dynamics and persistence of AAN-I-IFN specifically in MG patients. Thymoma probably makes a significant contribution to the development of AAN-I-IFN and independently increases the risk of hypoxemic COVID-19. This is consistent with previous reports of patients with thymoma and adverse reactions to the live attenuated vaccine against yellow fever and other severe viral illnesses, although AAN-I-IFN were not identified as the disease-causing agent at the time (*56, 57*). An increase in the expression of type I IFNs has been reported in the thymus of EOMG and MG-thymoma patients (*58, 59*), but with no increase in the type I IFN signature in blood (*60*). It would be interesting to test the correlation of this signature with the presence of AAN-I-IFN. Thymoma patients may also have other, currently undetected, auto-Abs or auto-reactive T cells.

Some MG patients with AAN-I-IFN suffered from mild COVID-19, indicating incomplete penetrance, as previously observed in other patients with AAN-I-IFN, including those with APS-1 (*28, 29, 61*). The penetrance of AAN-I-IFN is driven by the number and concentration of IFNs neutralized (*29*). Incomplete penetrance may be attributable to several factors: (i) differential avidity of auto-Abs for their target antigen, such that they are detected in our assay despite having limited physiological activity (*62*), (ii) host or viral genetic factors, (iii) age-related decline in immune function and (iv) overall immune competence, which may be further influenced by immunosuppressive therapies. In MG patients without detectable circulating AAN-I-IFN, the risk of severe COVID-19 pneumonia remains higher than in the general population, especially in MG patients with thymoma. This may be due to AAN-I-IFN not detectable in the blood but present in tissues (e.g., IgA or IgM auto-Abs (*63*)), AAN-I-IFN not detected by the assays available but nevertheless present and capable of neutralizing lower concentrations, immunosuppressive treatments, or the presence of other auto-Abs. Muscle weakness may also affect symptoms and disease severity. Overall, MG patients have a higher risk of hypoxemic COVID-19, especially if they have thymoma, and the risk may be further increased by the presence of AAN-I-IFN. We suspect that thymoma is directly involved in the production of AAN-I-IFN, which underlie hypoxemic COVID-19 pneumonia in these patients. Further studies are required to confirm this hypothesis and shed light on its mechanistic basis. Moreover, prospective cohort studies with baseline assessments of AAN-I-IFN levels are warranted to improve assessments of the risk of severe COVID-19 and other viral diseases conferred by AAN-I-IFN.

Our findings have several clinical implications. First, all MG patients should be screened for AAN-I-IFN, regardless of thymoma status, to identify those at higher risk of hypoxemic COVID-19 and potentially other severe viral infections that could be prevented, as MG is itself a risk factor for the development of such auto-Abs. Rapid blood assays could facilitate this screening (*64*). Second, these patients should be vaccinated against COVID-19 with mRNA vaccines and should receive booster doses, to reduce the risk of life-threatening COVID-19 (*45*), and they should also be vaccinated against influenza (*33*). Third, live-attenuated viral vaccines, such as YFV-17D, should be avoided (*41*). Fourth, these patients should avoid exposure to mosquitoes and ticks, given the very high risk of arboviral diseases in patients with AAN-I-IFN (*39, 40*). Fifth, unvaccinated MG patients of unknown auto-Ab status infected with SARS-CoV-2 should be monitored closely. The use of antiviral drugs and monoclonal antibodies against the virus is recommended^76–78^. In the absence of auto-Abs neutralizing IFN-β, treatment with IFN-β (*65*) may also be considered, although caution is required given the context of autoimmunity and the risk of immunization. It might be possible to use IFN-λ as an alternative (*66*). Other therapeutic strategies could be based on signaling-inert mutant IFN-I forms used as decoys to prevent IFN-I neutralization by auto-Abs (*62*), or on the use of chimeric autoantibody receptor T-cells (CAAR-T) specifically targeting AAN-I-IFN-producing B cells (*67*).

Several pathogenic aspects remain to be studied. Patients with APS-1 due to biallelic deleterious variants of *AIRE* develop multiple organ-specific autoimmune T-cell diseases and harbor AAN-I-IFN (*68, 69*), but they do not suffer from MG or have auto-Abs targeting the neuromuscular junction (*70*). By contrast, MG has been reported in a few patients with hypomorphic *RAG1* or *RAG2* variants, who typically present with AAN-I-IFN, and at least one patient with incontinentia pigmenti, a condition in which patients are known to harbor AAN-I-IFN (*71–75*). In patients with thymoma-associated MG, AAN-I-IFN may arise through a similar mechanism, in which an abnormal thymic architecture impairs the negative selection of IFN-I-specific autoreactive T cells. The resulting auto-Abs then neutralize the early IFN-I response to SARS-CoV-2 infection, thereby predisposing patients to life-threatening diseases. However, the molecular mechanisms and genetic causes driving the development of both MG and AAN-I-IFN in these patients have yet to be deciphered.

## Materials and Methods

### Patient recruitment

We recruited 171 patients with MG from several countries (France, Brazil, Tunisia, Spain, Canada, Switzerland) from whom samples were collected before the COVID-19 pandemic (*n*=86), during/after SARS-CoV-2 infection (confirmed by PCR and/or serology, *n*=85 hypoxemic or non-hypoxemic patients) including six patients from whom samples were collected both before and during/after infection, and two patients infected with SARS-CoV-2 and treated early. All analyses were performed in accordance with the relevant guidelines and regulations from the competent committees, and informed consent was obtained from all participants and/or their legal guardians. The details of the IRB/oversight body that provided approval or exemption for the research described are as follows: written informed consent for participation in this study was obtained from all participants and/or their legal guardians in accordance with local regulations, with approval from the institutional review board (IRB). For samples provided by the team (Sorbonne University/INSERM - UMRS974), investigations were approved by the local ethics committee (the relevant authorization numbers are ID RCB: 2006-A00164-47 (“Développement de modèles pour améliorer la prise en charge de la Myasthénie : de la connaissance fondamentale à l’application clinique (projet MYASTAID)”, which ethics approval was granted by the French Comité Consultatif de Protection des Personnes CPP IDF VII (Kremlin Bicêtre); and 2010-A00250-39 (“Construction d’une collection de tissus pour le projet FIGHT-MG”, which ethics approval was granted by the French Comité Consultatif de Protection des Personnes CPP IDF VI (Pitié Salpêtrière)). All the experiments and analyses were performed in accordance with the relevant guidelines and regulations. For the rest of the samples, ethics approval was granted by the French Comité Consultatif de Protection des Personnes - CCP - Ile-de-France VII, Kremlin Bicêtre, under protocols 2006-A00164-47 and 2010-A00250-39. Approval for the analysis of patient samples was also granted by the Institutional Review Boards at The Rockefeller University (New York, USA) and INSERM (France). We collected the serum or plasma samples obtained from these patients to test for the presence and activity of auto-Abs against type I IFNs. COVID-19 was classified as hypoxemic (severe or critical, requiring oxygen supplementation, *n*=48), or non-hypoxemic (mild/asymptomatic or moderate disease, not requiring oxygen supplementation, *n*=37).

### Immunoassays (ELISA, Gyros, Vidas)

ELISA was performed for IFN-α2a and IFN-ω, as previously described^84^. IgG levels were determined by Gyros, as previously described^58^. Anti-IFN-α2 IgG antibodies were quantified with the VIDAS anti-IFN-α2 IgG prototype assay (bioMérieux, Marcy L’Etoile, France), as previously described (*76*). This prototype had not been submitted to any regulatory agency for review at the time of writing. Concentrations are reported in ng/mL, and relative fluorescence values (RFV) are reported to obtain a numerical value for samples with a concentration of auto-Abs >5,000 ng/mL.

### Luciferase reporter assay

The ability of auto-Abs to neutralize IFN-α subtypes, IFN-β, or IFN-ω was determined with a reporter luciferase activity, as previously described^19^. Briefly, HEK293T cells were transfected with a firefly luciferase reporter plasmid containing 5 ISRE repeats, and with a *Renilla* luciferase plasmid. We stimulated the cells with IFNs (IFN-α2 or IFN-ω or IFN-β, at doses of 10 ng/mL and 100 pg/mL), diluted 1:10 in plasma from patients or controls, or the cells were left unstimulated in the presence of plasma. We measured the firefly luciferase signal, normalized against the *Renilla* luciferase signal. Finally, this ratio was normalized against the signal obtained in unstimulated conditions to calculate relative luciferase activity fold induction (RLA fold induction). Plates were washed three times in 0.005% PBTS between steps.

### IgG purification

IgG was purified on NAb Protein G Spin Columns (#89953, Thermo Fisher Scientific). Briefly, 100 µL plasma or serum was incubated with 400 µL Pierce Protein G IgG Binding Buffer (#21011, Thermo Fisher Scientific) and applied on the column. The columns were washed four times with 400 µL phosphate-buffered saline, and IgG was eluted with 400 µL 0.1 M glycine pH=2.7. The eluate was immediately neutralized with 40 µL Tris 1.5 M pH=8. Purified IgG samples were concentrated with Pierce Protein Concentrators PES, 50K MWCO (#88504, Thermo Fisher Scientific). The protein concentrations of the IgG-positive and IgG-negative fractions were determined with a Nanodrop 2000 spectrophotometer (Thermo Fisher Scientific).

### Protein array

Protein arrays (HuProt™ from CDI laboratories) were incubated in 5 mL blocking buffer, consisting of 2% bovine serum albumin and 0.05% Tween 20 in phosphate-buffered saline (PBS), for 90 min. The arrays were then incubated overnight in 5 mL blocking buffer per array with either blood donor serum or patient serum diluted 1:2000. Each array was then washed five times, for five minutes each, with 5 mL PBST (PBS + 0.05 % Tween 20). Alexa Fluor 647 goat anti-human IgG (Thermo Fisher Scientific Cat# A-21445, RRID:AB_2535862, LOT# 2286304) and Dylight® 550 goat anti-GST (Columbia Biosciences Corporation Cat# D9-1310, LOT# DY550011-18-003,) antibodies were diluted in blocking buffer (1:2000 and 1:10 000, respectively) and each array was incubated in 5 mL of the resulting mixture for 90 min. The arrays were washed five times, as described above. Incubations and washes were performed on an orbital shaker, with tubes wrapped in aluminum foil to block out light following the addition of fluorescent antibodies. Finally, each array was immersed three times in deionized water and centrifuged for approximately 30 seconds for drying. The arrays were scanned later the same day with an Innoscan 1100AL Fluorescence scanner (Innopsys) under the control of Mapix software and the resulting images were analyzed with GenePix Pro 5.1.0.19 and HuProt_v4.0.gal. Protein names are consistent with the aforementioned file in some cases, whereas proteins are named in accordance with HuProt_v4.0_Standard_Gal_Updated_04302019.gal in others. Data were normalized to compensate for the variation of signal intensity between experiments. All patient samples were screened in the same experiment, together with samples from two healthy controls. Data from additional healthy controls were included in separate protein array experiments. Signal intensities were extracted from the scanned image with GenePix Pro 5.1.0.19, with subtraction of the local background as follows:

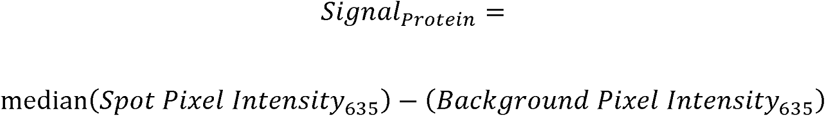

Each protein was printed in duplicate spots. The resulting signal for one sample is defined as follows:

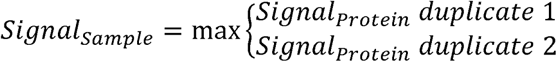

We screened for spurious results, by screening duplicates for large differences:

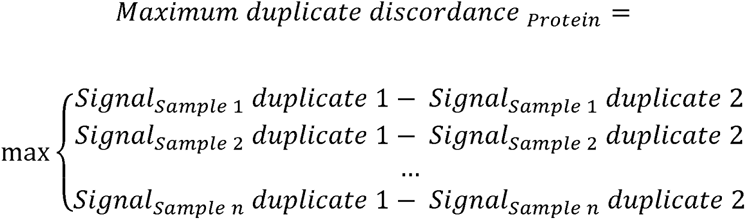

Mean signal intensity was calculated for case and control samples separately:

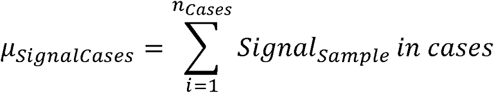

### Statistical analysis

ORs and *P* values for the effect of auto-Abs neutralizing each type I IFN in myasthenia gravis patients relative to healthy individuals from the general population, adjusted for age in three categories (≤50, 50-70, >70 years) and sex, were estimated by logistic regression analysis performed with the glm function of R software. OR and *P* values for the effect of auto-Abs, thymoma, thymectomy, rituximab or corticosteroid treatment in myasthenia gravis patients with hypoxemic pneumonia relative to patients with non-hypoxemic pneumonia, and OR and *P* values for the effect of auto-Abs in myasthenia gravis patients with thymoma relative to patients without thymoma, were estimated by Firth’s bias-corrected logistic regression adjusted for age, as implemented in the logistf package of R software. Within each analysis subset, Bonferroni correction was applied to account for multiple testing. For ease of interpretation, uncorrected *p*-values are reported throughout and it is indicated whether each finding remained statistically significant after Bonferroni correction. Where relevant, statistical test results are indicated in the corresponding figures. ns: not significant, \**P* < 0.05, \*\**P* < 0.01, \*\*\**P* < 0.001, \*\*\*\**P* < 0.0001.

### Study approval

Written informed consent for participation in this study was obtained from all participants and/or their legal guardians in accordance with local regulations, with approval from the institutional review board (IRB). For samples provided by the team (Sorbonne University/INSERM – UMRS974), the investigations were approved by the local ethics committee (relevant authorization numbers: ID RCB: 2006-A00164-47 and 2010-A00250-39). All the experiments and analyses were performed in accordance with the relevant guidelines and regulations.

## Data Availability

All the data required to evaluate the conclusions in the paper are available in the main text.

## Author Contributions

A. G., A. Co., J.-L. C., A. P., P. B., and E. J. designed the research. A. G., T. L. V., J. R., Q. P., L. B., J. P., A. Ce., M. M., S. P., and K. S. conducted experiments and acquired data. A. G., A. Mar., A. Mai., T. L. V., J. R., Q. P., L. B., J. P., A. Ce., N.L, and A. Co. analyzed data. P. L., M. A., C. G., C. Bi., R. L. L., S. S., C. T., A. N. P., A.-M. G., N. H., D. A., A. V., S. A., K. B., K. H., L. K., R.-N. V.-Q., C. C., M.-C. F., E. R., G. N., E. D. P. E., M. D. P. A. M., E. Z., Z. S., A. Ra., A. Ri., S. Bel., S. Ben. S., A. H., E. F., I. D., C. Ba., N. G., V. B., S. D., F. T., R. F., F. A., F. W., L. A., T. M., M.-A. A., R. L. P., and G. S. recruited patients and collected clinical information. A. G., J.-L. C., A. P., P. B., and E. J. wrote the initial manuscript. All the authors edited the manuscript.

## Data sharing plan

All data needed to evaluate the conclusions in the paper are available in the main text.

## Acknowledgments

We thank the patients and their families for placing their trust in us. We thank the members of both branches of the Laboratory of Human Genetics of Infectious Diseases. We thank Y. Nemirovskaya, M. Woollett, D. Liu, S. Boucherit, A. Geraldo, G. Goutbi, M. Chrabieh, and L. Lorenzo for administrative assistance. We also thank the staff of the Imagine facilities: C. Bureau, L. Colonna, S. Paillet, N. Ghouas, and M. Sy. We also thank the legal team and technology transfer staff of the Imagine Institute: E. Rubino, W. Loewen, D. Beudin, and N. Wuylens. We thank all the staff of the Imagine Institute, Necker Hospital, and Necker sorting center for help.

---

**Figure S1.**
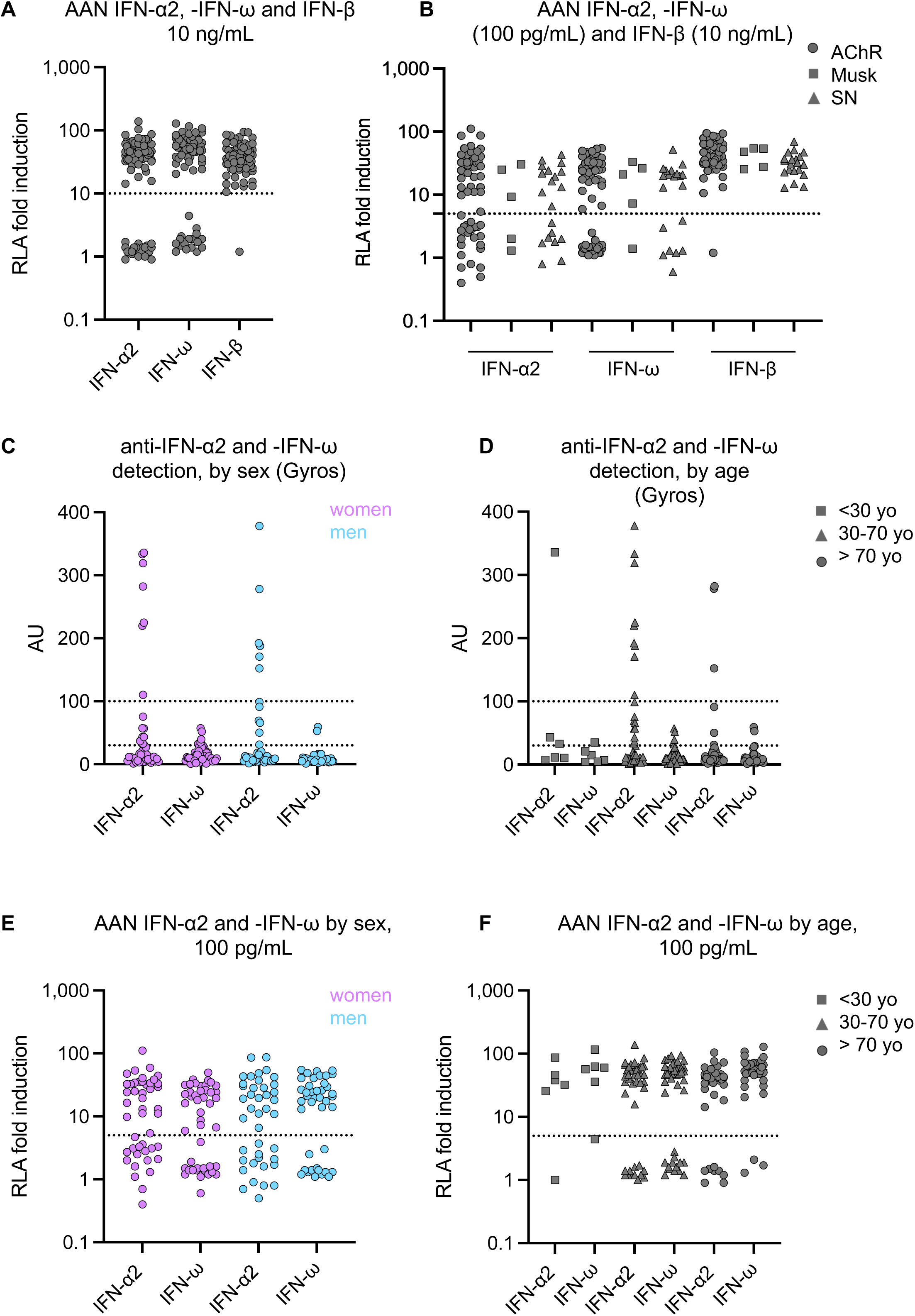

**Figure S2.**
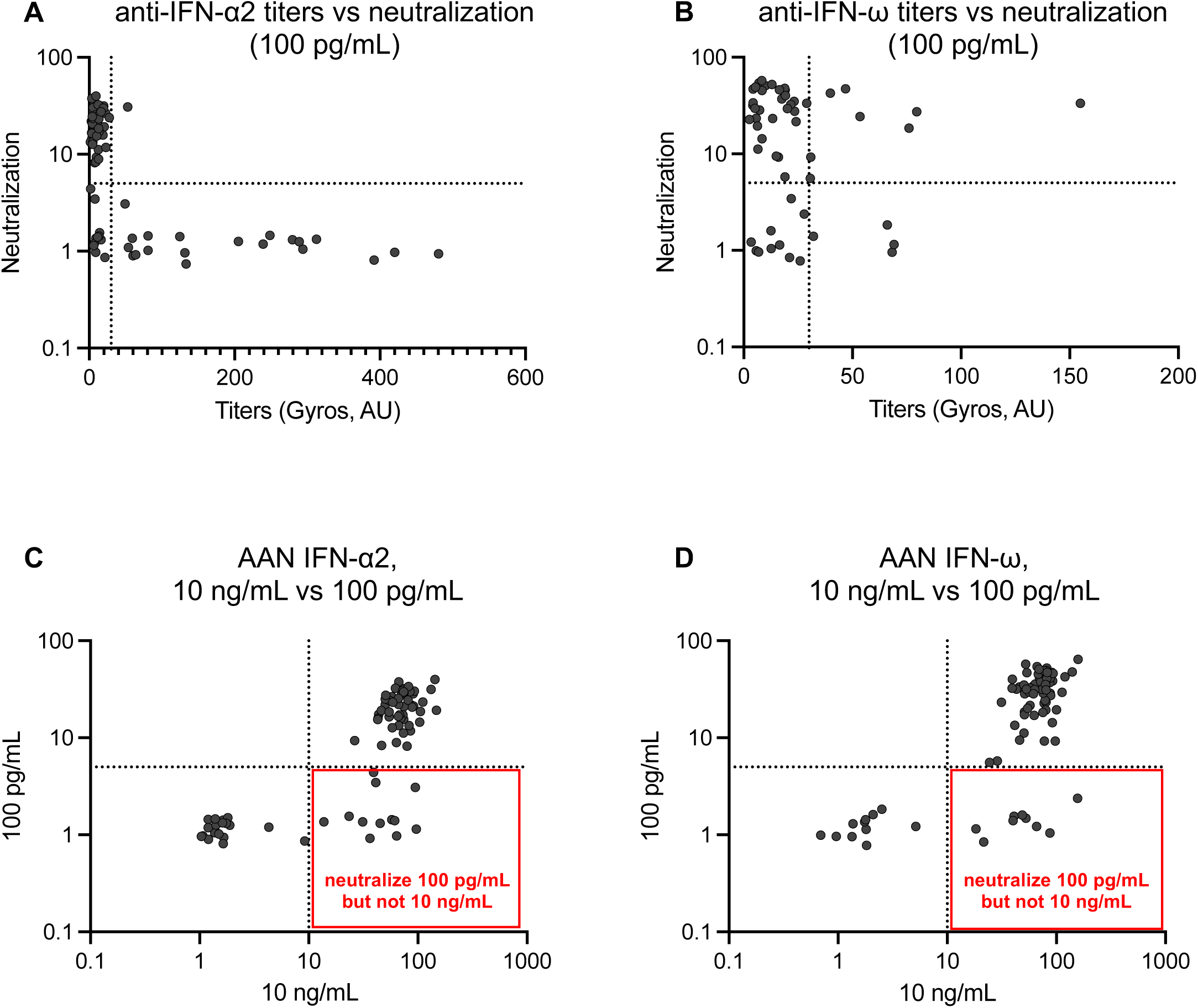

**Figure S3.**
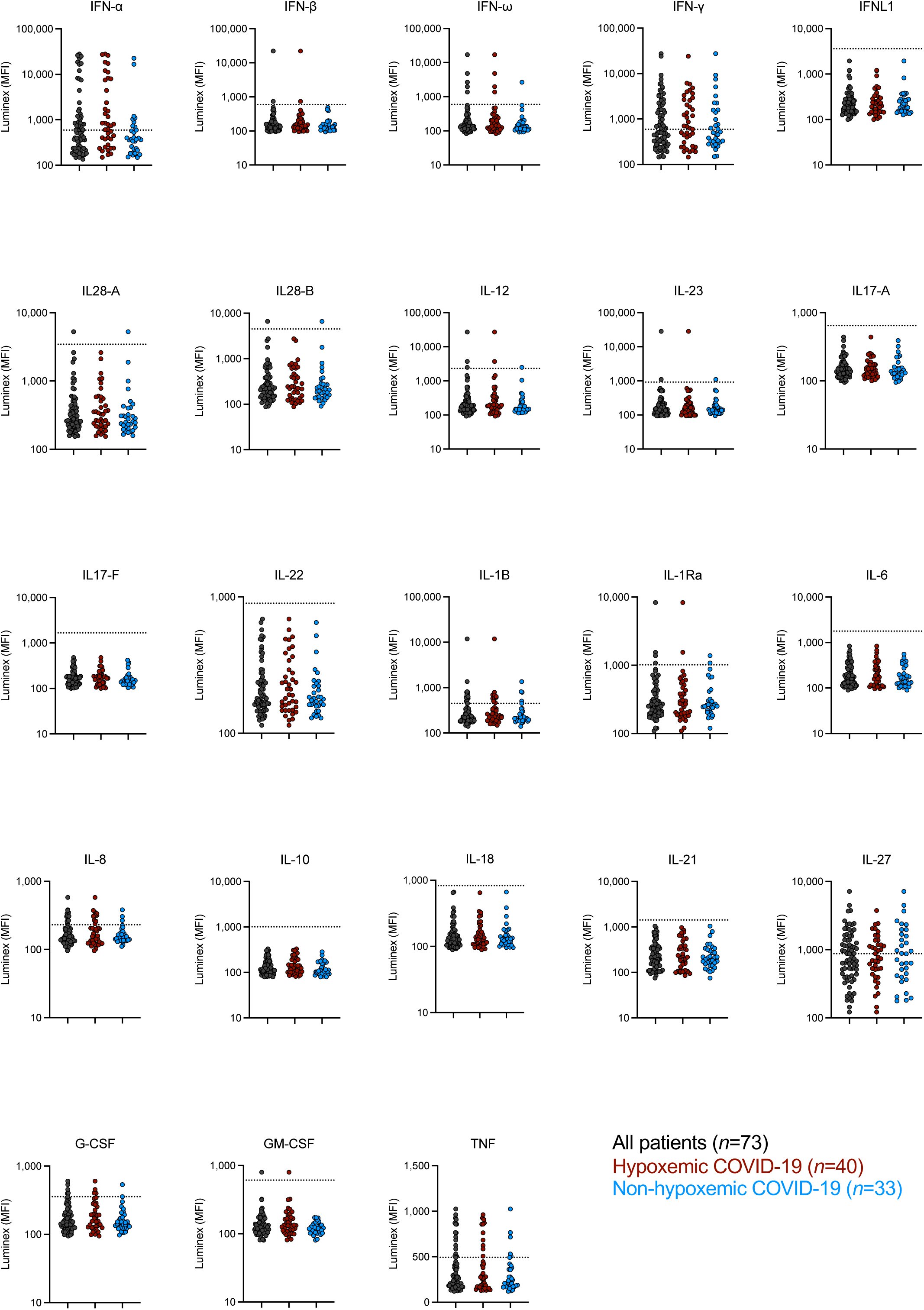

**Figure S4.**
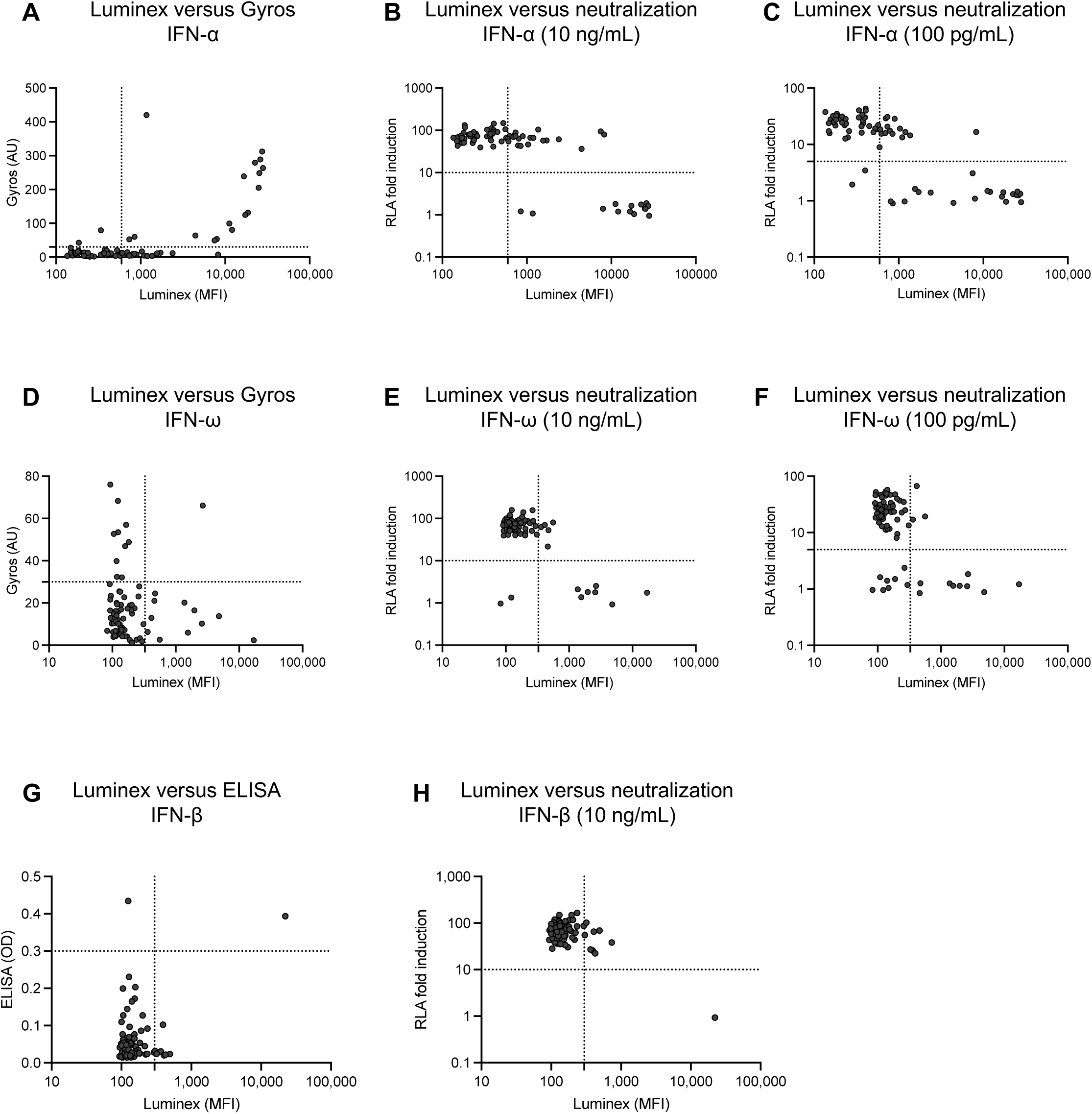

