## Supplemental material for "High risk of hypoxemic COVID-19 pneumonia in myasthenia gravis patients with type I IFN autoantibodies"

PNAS

**Figure S1**

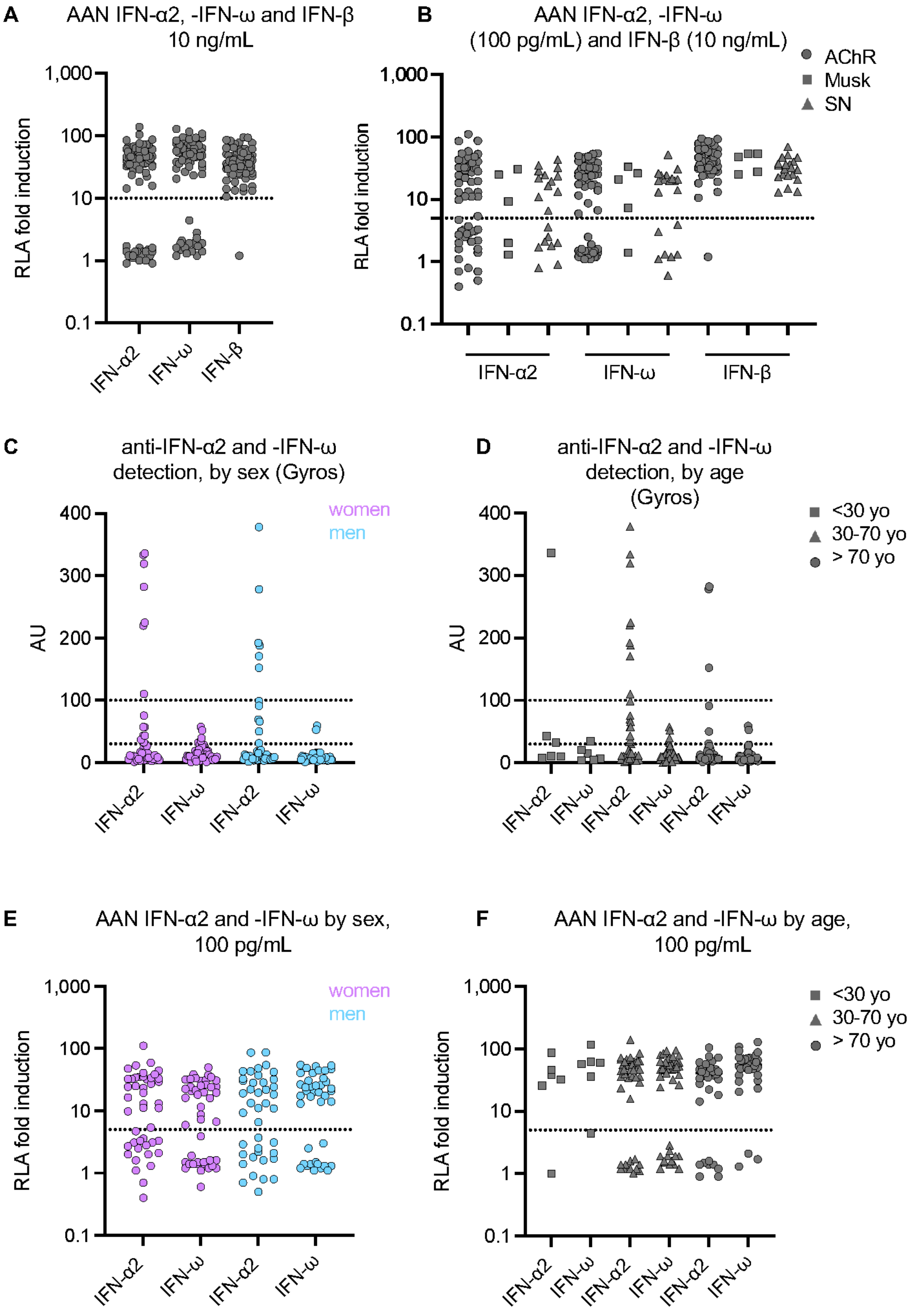


**Figure S1: Auto-Abs against type I IFNs in the 86 MG patients before the episode of COVID-19, by age, sex or MG serotype**

**(A)** AAN-I-IFNs neutralizing 10 ng/mL IFNs. **(B)** AAN-I-IFNs by MG serotype. **(C)** Detection by Gyros of auto-Abs against IFN-α2 and IFN-ω, by sex. **(D)** Detection by Gyros of auto-Abs against IFN-α2 and IFN-ω, by age group. **(E)** AAN-IFN-I, by sex. **(F)** AAN-IFN-I, by age group.

**Figure S2**

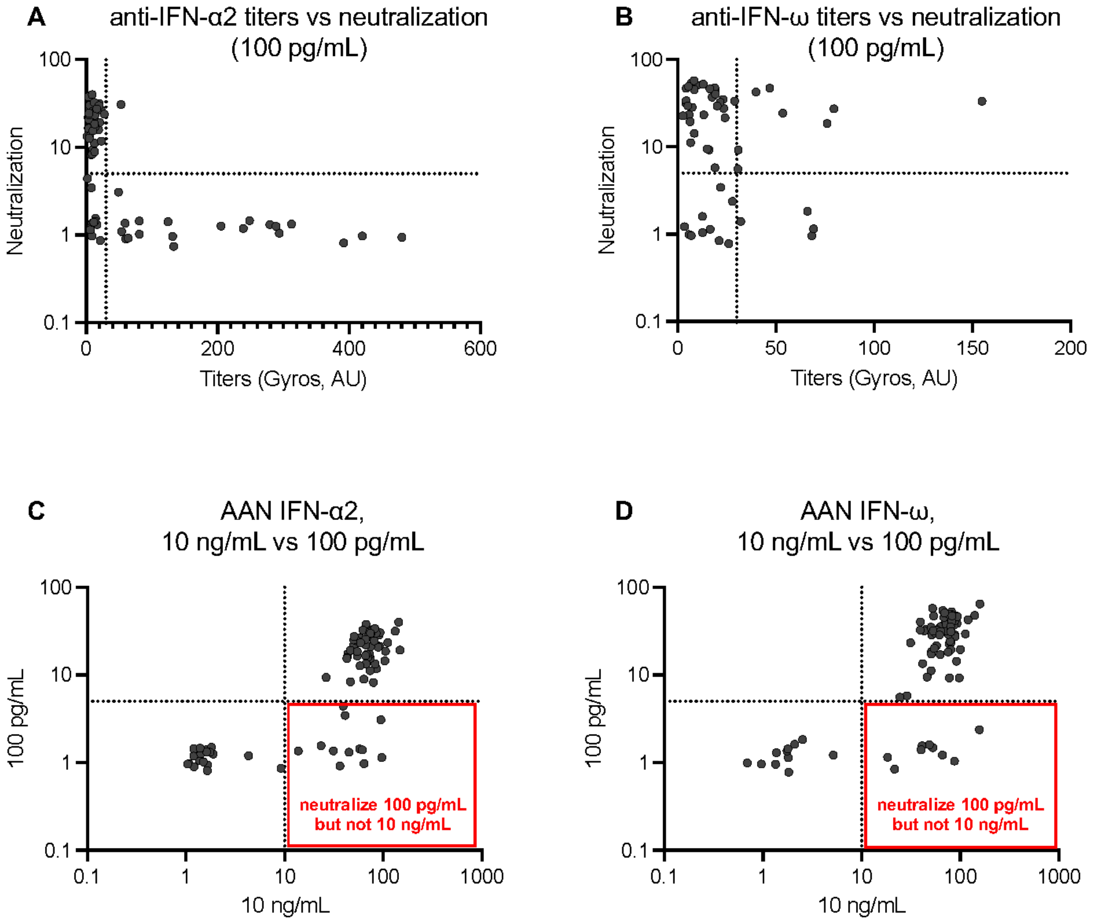


**Figure S2: Correlation of neutralization data with auto-Ab titers and correlation of neutralization between high and low IFN concentrations**

**(A)** Correlation between IFN-α2 titers determined by Gyros and neutralization of 100 pg/mL IFN as assessed by luciferase assay. **(B)** Correlation between IFN-ω titers determined by Gyros and neutralization of 100 pg/mL IFN as assessed by luciferase assay. **(C)** Correlation between the neutralization of IFN-α2 at concentrations of 100 pg/mL and 10 ng/mL, as assessed by luciferase assay. **(D)** Correlation between the neutralization of IFN-ω at concentrations of 100 pg/mL and 10 ng/mL, as assessed by luciferase assay.

**Figure S3**

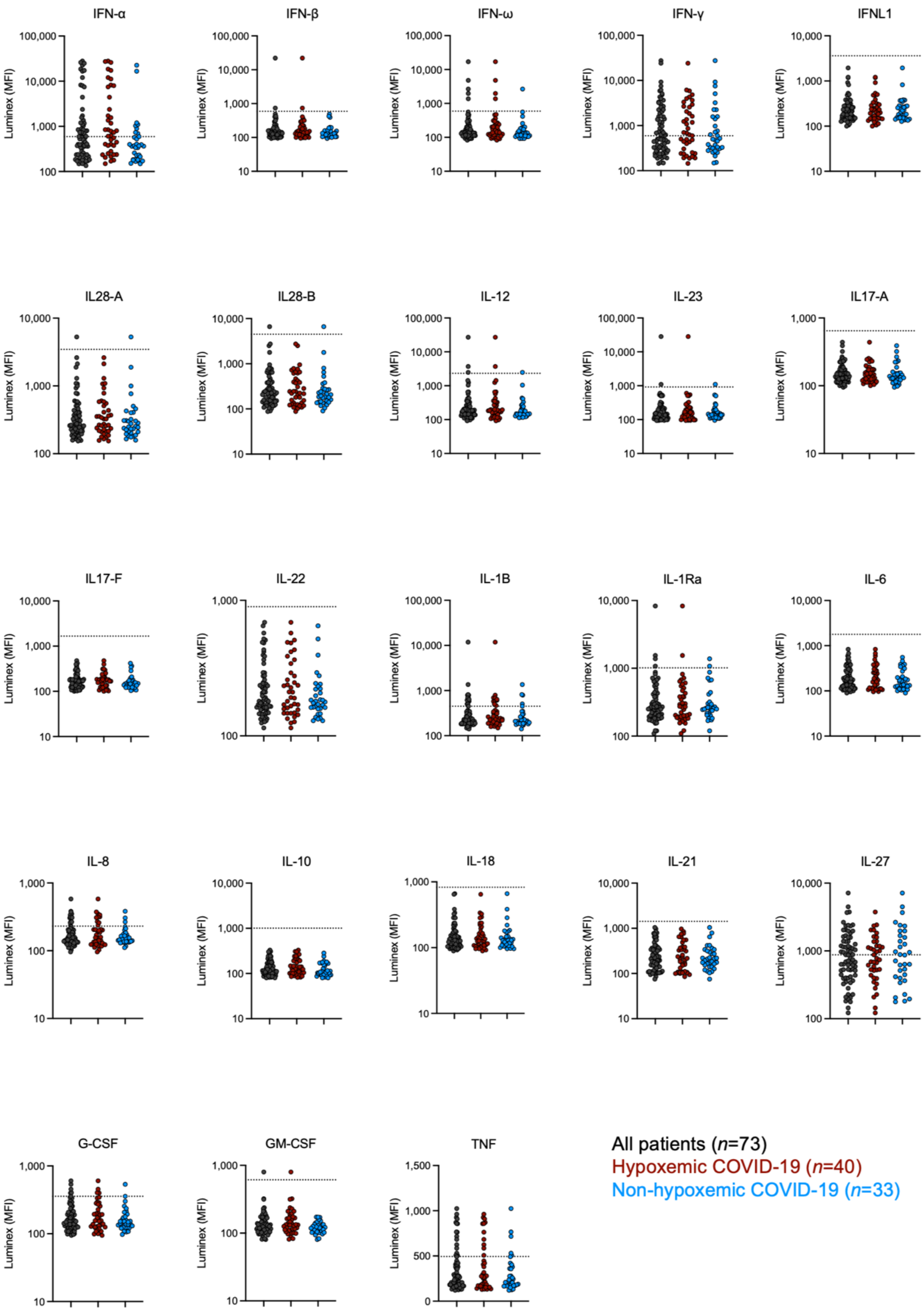


**Figure S3: Detection of a panel of auto-Abs against cytokines by Luminex® assays**

The threshold was defined as four standard deviations above the mean signal for healthy donors.

### Figure S4

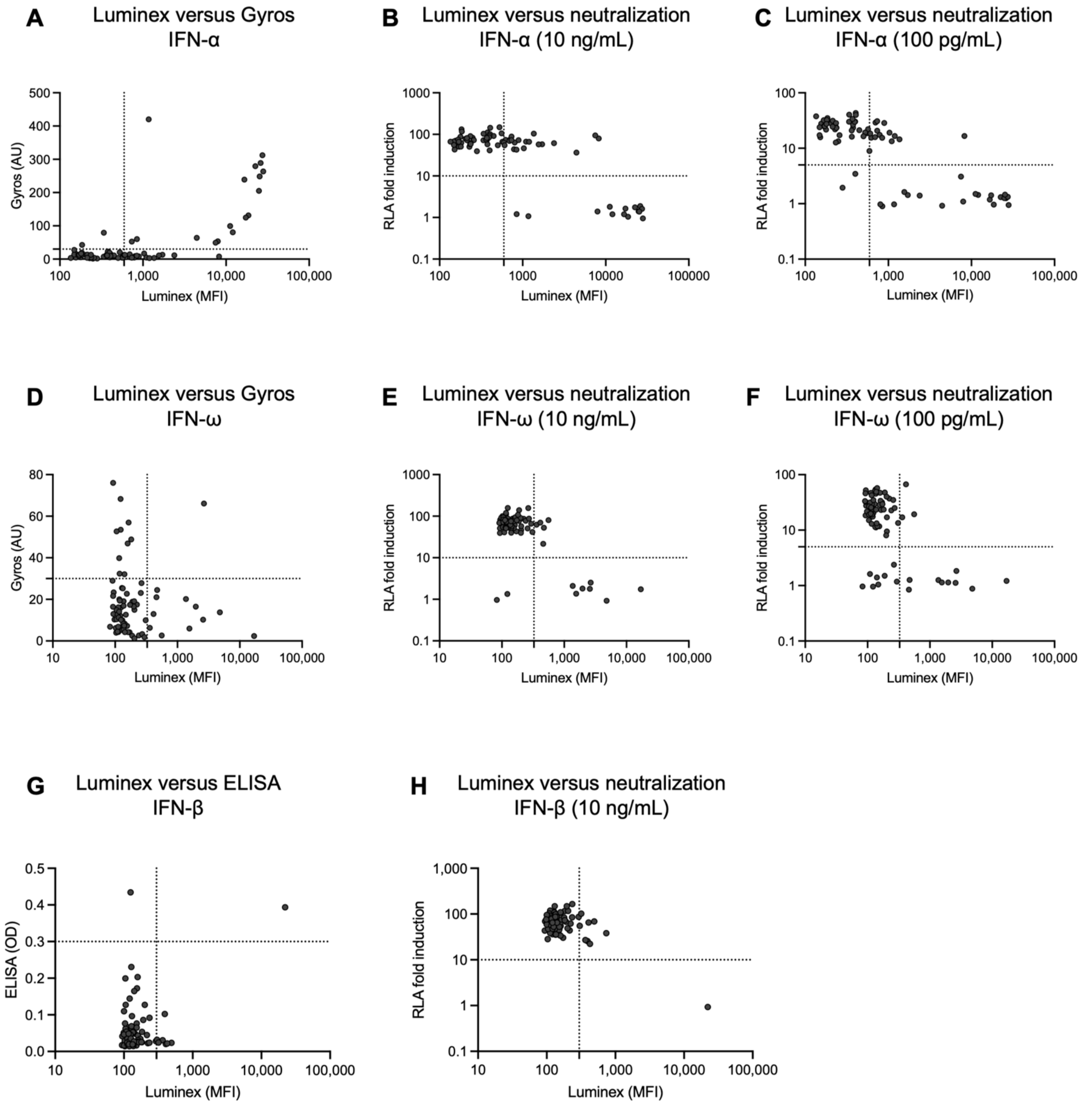


**Figure S4: Correlation of Luminex data with Gyros data and/or neutralization data for auto-Abs against IFN-α2, IFN-ω and IFN-β.**
